# Short tandem repeats significantly contribute to the genetic architecture of metabolic and sensory age-related hearing loss phenotypes

**DOI:** 10.64898/2026.02.17.26346449

**Authors:** Samah Ahmed, Kenneth I. Vaden, Judy R. Dubno, Galen E.B. Wright, Britt I. Drögemöller

## Abstract

Age-related hearing loss (ARHL) is a progressive, bilateral decline in hearing ability that affects one in four individuals over 60 years of age worldwide. While previous genome-wide association studies (GWAS) have identified distinct single-nucleotide variants (SNVs) associated with metabolic and sensory ARHL phenotypes, the contribution of short tandem repeats (STRs) – a neglected yet important class of genetic variants – remains poorly understood. To address this gap, TRTools was used to impute STRs from a high quality, sequencing-derived SNV-STR reference panel to investigate the association between STRs and metabolic and sensory estimates. Heritability analyses revealed that while STRs contribute to estimates of both ARHL components, this class of variation plays a more important role in metabolic hearing loss (6%), which typically increases with age, compared to sensory hearing loss (4%). Further, the inclusion of this class of variant into GWAS analyses uncovered an association between a haplotype consisting of two missense variants (rs7714670 and rs6453022) and an intronic STR (chr5:73778077:A_16_) in *ARHGEF28* (*P*=3.30×10^-9^), proving further insight into the variants driving this previously identified signal. Notably, burden analyses revealed that rare and longer repeats were associated with an increased risk of the metabolic phenotype and a reduced risk of the sensory phenotype. Functional annotation of significant and nominally significant STRs revealed potential effects on gene expression and splicing of nearby genes. Our findings provide the first evidence that STRs explain some of the missing heritability of ARHL phenotypes and create an STR resource for researchers to use in future analyses.

## 1. Introduction

Age-related hearing loss (ARHL) is one of the most common sensory deficits in older adults, characterized by a gradual, bilateral decline in hearing sensitivity that typically manifests later in life.^1^ According to the World Health Organization, one in four individuals over 60 years of age is affected by ARHL worldwide.^2^ As the aging population is projected to nearly double in the coming decades, the number of individuals affected by ARHL is expected to rise substantially, placing a significant burden on the healthcare system.^3^ This highlights the need for new preventive and therapeutic interventions.

ARHL has a strong genetic component, with inherited variation playing a key role in determining individual susceptibility and influencing the progression and severity of hearing loss.^4,5^ Notably, ARHL is not a single, uniform phenotype. Instead, it reflects a spectrum of cochlear and other auditory pathologies that are often not adequately captured by self-reported hearing loss questionnaires. To better account for this heterogeneity, classifying ARHL into two primary phenotypes based on audiometric patterns has been proposed: metabolic hearing loss, associated with degeneration of the stria vascularis and associated reduction in the endocochlear potential, and sensory hearing loss, resulting from dysfunction of and damage to outer hair cells.^6–8^ These phenotypes likely arise from distinct biological mechanisms and, accordingly, may have different genetic risk factors.

Genome-wide association studies (GWAS) support this phenotypic distinction. For example, our recent GWAS analyses identified distinct genetic variants associated with each phenotype. A missense variant in *ARHGEF28* was associated with the metabolic phenotype, while a missense variant in *KLHDC7B* was linked to the sensory phenotype.^9^ These findings suggest that phenotypic refinement enhances the power of genetic analyses, aiding in the confirmation of biological mechanisms underlying ARHL and its subtypes. However, a major limitation of conventional GWAS is that these studies mainly focus on single-nucleotide variants (SNVs), due to past technological and bioinformatic constraints, thus missing other important forms of genetic variation that contribute significantly to human traits and diseases.^10^

Short tandem repeats (STRs) are repetitive DNA sequences typically composed of motifs ranging from one to six base pairs.^11^ STRs occur at over one million loci across the genome, collectively accounting for approximately 3% of the human genome.^12^ These elements are highly dynamic, exhibiting frequent gain or loss of repeat units during DNA replication, with mutation rates up to 100 million times higher than those of SNVs.^12^ While STR expansions are well known to cause Mendelian disorders such as Huntington disease and Fragile X syndrome,^13^ emerging evidence suggests that they also influence susceptibility to complex neurological traits, including Alzheimer’s disease.^14^ Notably, the vast majority of STRs (i.e., approximately 92%) are located in non-coding regions of the genome^15^ where they can impact gene expression, DNA methylation, and other regulatory mechanisms that represent important biological processes that contribute to complex polygenic traits.^16–18^

STRs pose unique challenges for genotyping, as they cannot be directly assayed with the traditional DNA-array technologies used in GWAS. However, recent advancements in whole-genome sequencing have enabled the development of comprehensive SNV-STR reference panels.^19,20^ Coupled with the development of tandem repeat-aware bioinformatics tools, these advances now allow STRs to be reliably imputed from SNV genotype data and incorporated into GWAS of complex traits.^21^ Given the late-onset nature of ARHL, STRs may contribute to the etiology of this phenotype through mechanisms such as age-dependent gene dysregulation (e.g., driven by somatic repeat expansion), as has been observed in other neurodegenerative and sensory disorders.^22,23^

Here, we assess the potential role of STRs in the genetic architecture of metabolic and sensory components of ARHL. We leverage a large-scale GWAS of 1.2 million STRs, imputed from SNV array data, from the Canadian Longitudinal Study on Aging (CLSA). Our study identifies, for the first time, the contribution of STRs to specific estimates of metabolic and sensory components of ARHL. Exploring the association between STRs and ARHL can enhance our understanding of the genetic factors that contribute to this common age-related condition.

## 2. Methods

### 2.1. Genotype and phenotype data

The genome-wide SNV genotype, audiologic and clinical/demographic data used in this study, along with all associated processing steps, have been previously described in detail by Ahmed *et al*.^9^ In brief, genomic and audiologic data were obtained from healthy older individuals participating in the CLSA.^24^ Marker- and sample-level quality control (QC) were conducted following the CLSA workflow.^25^ Briefly, genotype markers with a genotype missingness rate > 0.05, minor allele frequency (MAF) < 0.01, Hardy–Weinberg equilibrium (HWE) *P* value < 1×10^-6^, or imputation quality (R^2^) ≤ 0.8 were excluded. Related individuals (pairwise kinship coefficient > 0.125), as well as those exhibiting extreme heterozygosity, high genotype missingness, or discordance between genetic and self-reported sex, were also excluded. Estimates of metabolic and sensory components of ARHL were determined using the method described by Vaden *et al*.,^6^ which derives these estimates from the audiogram of the better hearing ear. After stringent filtering and preprocessing, a total of 18,985 samples and 8,053,152 markers were included in downstream analyses. All analyses were conducted using the genome build GRCh38.

### 2.2. Imputation of STRs

To perform STR imputation, we used the SNV-STR reference panel consisting of 3,202 samples from the 1000 Genomes Project.^20^ This panel included high-coverage (30x) whole-genome sequences, generated using Illumina NovaSeq 6000 instruments, from 603 trios from 5 superpopulations: African (*n*=893), European (*n*=633), East Asian (*n*=601), South Asian (*n*=585) and American (*n*=490).^26^ A total of 1,070,762 STRs were genotyped by Ziaei Jam *et al*. using four different genotypers (HipSTR, GangSTR, ExpansionHunter and adVNTR) to capture diverse repeat types. Calls from the four genotypers were combined using EnsembleTR.^20^ Using these data, we imputed STRs for autosomal chromosomes using Beagle v5.5.^27,28^ STRs with low imputation quality (DR² < 0.7) were excluded using BCFtools.^29^

### 2.3. Heritability testing

We estimated the contribution of STRs to the heritability (*h^2^*) of metabolic and sensory estimates of ARHL by comparing STR *h^2^* to SNV *h^2^* using linkage disequilibrium (LD)- and minor allele frequency (MAF)-stratified GREML (GREML-LDMS). This tool was used as it is suited to estimate *h^2^* from imputed variants and account for LD structure by stratifying SNVs into four quartile bins based on the distribution of individual SNV LD scores.^30,31^ The following formula was used to calculate STR *h^2^* relative to SNV *h^2^* :

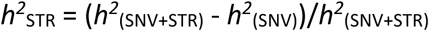

### 2.4. Genome-wide association testing

We performed genome-wide association testing of STRs with rank-transformed metabolic and sensory estimates using the open-source TRTools package. This enabled filtering, QC and analysis of multi-allelic STRs obtained from different genotyping tools.^21,28^ Clinical and demographic variables, along with the first ten genetic principal components (PCs) described in our previous GWAS by Ahmed *et al*. were also included as covariates in these analyses to adjust for non-genetic confounders and population stratification.^9^ This includes age, sex (for the sensory), hypertension, diabetes and the opposite hearing loss phenotype. Of the original 18,985 samples, 18 (0.09%) were removed due to missing covariates, leaving a final dataset of 18,967 samples (**Supplementary Figure 1**).

As Beagle STR imputation does not retain key STR-specific format fields (e.g., repeat motifs, allele lengths), we used annotaTR from TRTools to restore these attributes from the reference panel described above. This ensured compatibility with additional downstream analyses performed with TRTools.^21^ The quality of imputed STRs was summarized using qcSTR through TRTools (**Supplementary Figure 2**). Next, association testing was performed using associaTR, which tested the association between metabolic and sensory ARHL estimates and the sum of STR allele-length dosages across both chromosomes.^7,11^ A Bonferroni-corrected threshold of *P*<1.5×10^-7^ was considered genome-wide significant, and *P*<1×10^-5^ was considered suggestively significant in all STR-based analyses.^30^ Findings from the STR-based GWAS were compared to our previously published SNV-based GWAS of the same cohort to identify distinct and overlapping loci associated with each phenotype.^9^

### 2.5. Fine-mapping of STR signals

To identify likely causal STRs within genome-wide significant regions, we applied fine-mapping using the Sum of Single Effects (SuSiE) model (v0.14.2) using the susieR package in R.^33^ For each genome-wide significant region, we included variants located within ±1 Mb of the top STR signal. Fine-mapping was conducted across both STRs and SNVs in the region to identify the most likely causal variants. Posterior inclusion probabilities (PIPs) were used to estimate the likelihood of each variant being causal, with PIP>0.3 considered as evidence for potential causality, as previously described by Cui *et al*.^34^

To determine whether ARHL-associated STRs and SNVs occurring within the same locus reflected independent signals, we performed reciprocal conditional analyses. Specifically, we evaluated the association of the top metabolic ARHL-associated STR (i.e., intronic STR chr5:73782133:A_n_), while conditioning on the lead SNV (i.e., rs6453022; p.Pro284Gln) from our previous GWAS, and conversely, assessed the SNV association while conditioning on the STR. For each conditional analysis, the conditioning variant was included as a covariate in a linear regression model.

Further, we performed haplotype analysis using the GHap R package to evaluate the combined effects of the STR identified as most likely to be causal within the locus on chromosome 5 (i.e., intronic STR chr5:73778077:A_n_) and nearby missense SNVs (i.e., rs6453022; p.Pro284Gln and rs7714670; p.Trp225Arg) identified from our previous GWAS.^35^ Associations were then tested using PLINK v1.9, excluding haploblocks with MAF<0.01.^36,37^

### 2.6. STR functional annotation

To investigate the predicted functional impact of STRs on the genome, all variants reaching suggestive genome-wide significance (*P*<1×10^-5^) were mapped to genes using GENCODE v47.^38^ In addition, these STRs were annotated using AlphaGenome, a recently developed deep learning model trained on large multi-omic datasets from consortia such as ENCODE, FANTOM5 and GTEx. AlphaGenome predicts the effects of genetic variants on molecular phenotypes, including gene expression, chromatin accessibility, and splicing patterns.^39^ We focused on prioritized STRs with MAF>0.01, analyzing their predicted effects within a 100 kb window in brain tissue (ontology ID: UBERON:0000955), which serves as a relatively close proxy for the inner ear, to assess the effect of STRs on various molecular brain phenotypes across multiple brain regions.^34,40^ STRs with a quantile score>0.95 (representing variants with predicted impacts ranking in the top 5% compared to a background distribution of ∼300,000 common variants) were considered to have a potentially strong predicted functional impact.

Further, these annotations were supplemented using the brain TR-xQTLs atlas (https://wlcb.oit.uci.edu/TRxQTL).^34^ This STR atlas integrates data from three amyotrophic lateral sclerosis (ALS) and Alzheimer’s disease cohorts and includes gene expression quantitative trait loci (eQTL), splicing quantitative trait loci (sQTL), DNA methylation quantitative trait loci (mQTL), chromatin accessibility quantitative trait loci (hQTL), protein expression quantitative trait loci (pQTL), H3K9ac histone modifications, and alternative polyadenylation (APA) data. Repeat classes of identified STRs were annotated using RepeatMasker implemented through UCSC Genome Browser.^41^ These mQTLs were annotated using Illumina Human Methylation EPIC v2.0 manifest https://github.com/zhou-lab/KYCG_knowledgebase_EPICv2.^42^

### 2.7. Burden of STR expansions

To investigate the cumulative burden of STR repeat expansions on the risk of ARHL metabolic and sensory estimates, we identified STRs groups with repeats ≥1, ≥5, ≥10 and ≥20 longer than the reference at frequency cut-offs ≤5, ≤10, ≤100 and no cut-off.^43^ We calculated the total number of repeats per individual for each group. We then conducted multiple linear regressions to examine the association between the sum of repeats and the rank-transformed sensory and metabolic ARHL estimates. FDR corrected *P* values<0.05 were considered significant.

## 3. Results

### 3.1. Contribution of STRs to the heritability of metabolic and sensory components of ARHL

Following imputation, 262,839 STRs with low imputation quality (DR²<0.7) were excluded, resulting in 807,923 high-quality STRs remaining for GWAS analysis (**Supplementary Figure 1**). Using these data, we estimated the *h^2^* for the estimates of metabolic and sensory components of ARHL using models that incorporated SNVs alone or in combination with STRs. For the metabolic estimates, the SNV-only model yielded an *h^2^* of 0.137 (*P*=1.6×10^-3^), while the SNV+STR model showed a slightly higher *h^2^* of 0.146 (*P*=1.33×10^-3^), indicating that STRs contribute approximately 6.16% to the total *h^2^* of metabolic ARHL. For the sensory estimates, the SNV-only model revealed slightly higher *h^2^* estimates when compared to metabolic ARHL (*h^2^*=0.154; *P*=2.65×10^-3^). However, examination of the combined SNV+STR model (*h^2^*=0.160; *P*=3.89×10^-3^) revealed that while STRs still contribute to the *h^2^* of sensory ARHL, their contribution is smaller than what is observed for metabolic ARHL (3.75% contribution).

### 3.2. STRs are significantly associated with metabolic and sensory hearing loss

Genome-wide association analyses were conducted separately for metabolic and sensory estimates in 18,967 healthy aging participants from the CLSA cohort. No STRs reached genome-wide significance for the sensory phenotype. For the metabolic phenotype, our analysis identified four STRs reaching genome-wide significance (*P*<1.49×10^-7^), all mapping to *ARHGEF28* on chromosome 5 (**Figure 1**, **Table 1, Supplementary Tables 1 and 2**). Previous investigation of SNVs in this region by Ahmed *et al.* also identified two predicted benign *ARHGEF28* missense SNVs in this locus, rs6453022 (p.Pro284Gln) and rs7714670 (p.Trp225Arg) (*P*<5×10^-8^).^9^

**Figure 1:**
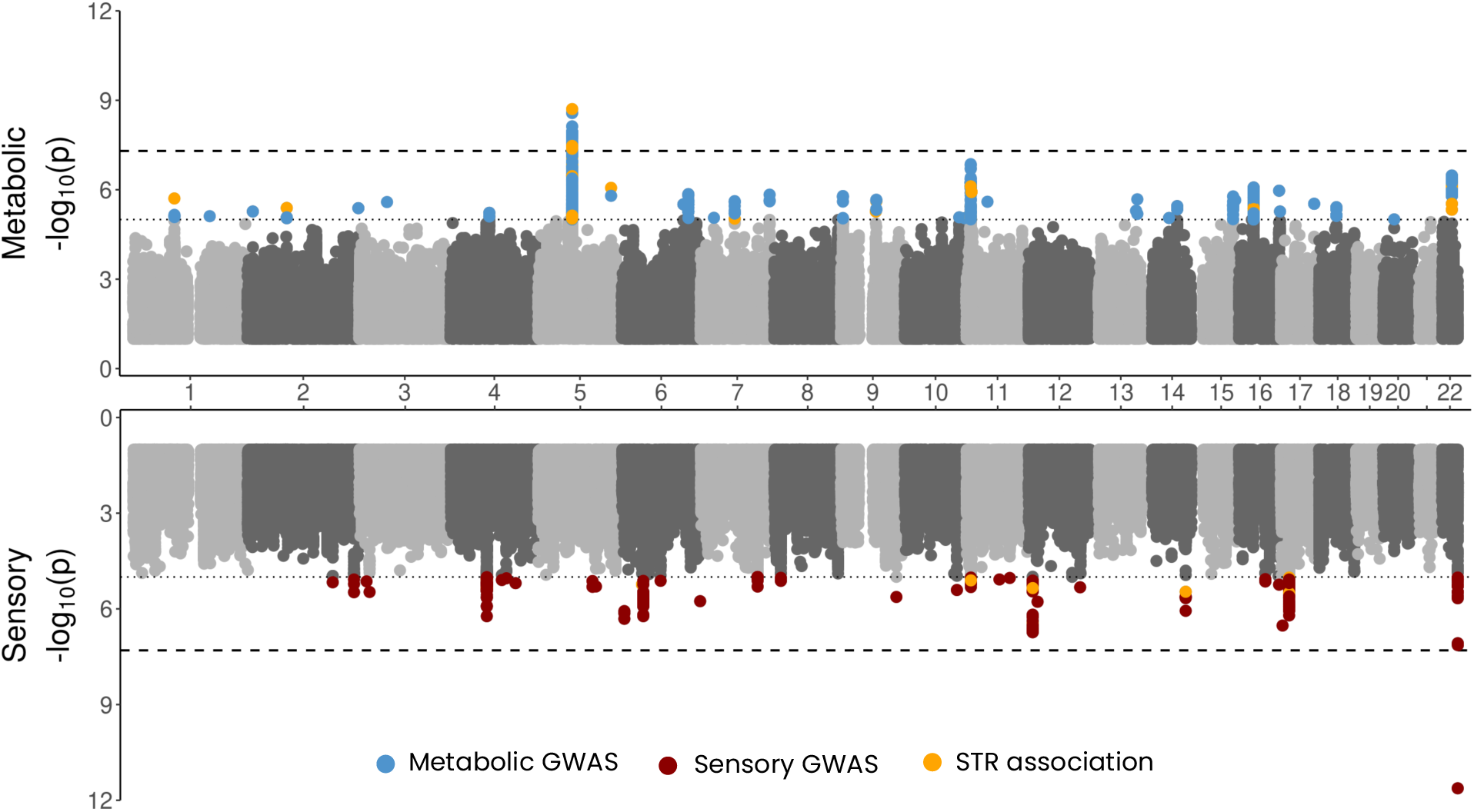
Association of STRs with metabolic and sensory phenotypes overlaid on SNV GWAS results. Blue and red points represent SNVs that reach a suggestive level of significance (*P*=1x10^-5^) for the GWAS for metabolic and sensory phenotypes in the upper and lower panel, respectively. Orange points represent STRs that reach a suggestive level of significance. STRs: Short tandem repeats; SNV: Single-nucleotide variants; GWAS: Genome-wide association study. The dotted horizontal lines indicate the suggestive threshold (*P*=1x10^-5^) and the dashed horizontal lines indicate the significance threshold (*P*=1.49x10^-7^).

**Table 1:** Summary of lead and fine-mapped SNVs and STRs at the locus significantly associated with the metabolic phenotype.

| CHR | Position<br>(rsID; amino acid<br>change) | STR motif/SNV<br>nucleotide change | STR reference<br>allele repeat<br>length | STR alternative<br>alleles repeat length | GWAS *P* value | GWAS<br>standardized<br>beta | Combined STR +<br>SNV PIP score | Mapped<br>gene |
| --- | --- | --- | --- | --- | --- | --- | --- | --- |
| 5 | 73782133 | A<sub>n</sub> | 16 | 15, 16, 17, 18, 19,<br>20, 21 | 1.97×10<sup>-9</sup> | 0.29 | 0.011 | ARHGEF28 |
|  | 73780643 | T<sub>n</sub> | 17 | 16, 17, 18 | 3.38×10<sup>-8</sup> | -0.80 | 0.003 |  |
|  | 73793799 | T<sub>n</sub> | 16 | 13, 14, 15, 16, 17 | 4.22×10<sup>-8</sup> | -0.41 | 0.026 |  |
|  | 73754888 | ATTT<sub>n</sub> | 5 | 4, 5, 6 | 8.24×10<sup>-8</sup> | -0.86 | 0.038 |  |
|  | 73778077 | A<sub>n</sub> | 16 | 15, 17, 18, 19 | 4.54×10<sup>-5</sup> | 0.41 | 0.063 |  |
|  | 73780686<br>(rs6453022;<br>p.Pro284Gln) | C>A | - | - | 2.67×10<sup>-9</sup>* | -2.58* | 0.029 |  |
|  | 73776529<br>(rs7714670;<br>p.Trp225Arg) | T>C | - | - | 2.51×10<sup>-8</sup>* | 2.43* | 0.006 |  |

CHR: Chromosome, STR: Short Tandem Repeats, SNV: Single Nucleotide Variant, GWAS: Genome-wide association study, PIP: Posterior Inclusion Probability.
\*Results from Ahmed *et al*<sup>9</sup>

### 3.3. Fine-mapping, conditional analyses and annotation of the top STR signals

Given the high LD between the SNVs and STRs that were significantly associated with metabolic ARHL (**Figure 1**), we conducted reciprocal conditional analyses on the lead STR (chr5:73782133:A_n_) and lead SNV (rs6453022; p.Pro284Gln) identified from the GWAS. These analyses revealed that the STR association was attenuated after conditioning on the lead GWAS SNV (*P*=0.10 vs. unconditioned *P*=1.97×10^-9^), suggesting that the SNV (rs6453022; p.Pro284Gln) explains a substantial portion of the observed signal. Conversely, conditioning the SNV association on the lead STR also reduced the significance of the SNV association (*P*=6.41×10^-4^ vs. unconditioned *P*=2.67×10^-9^), but to a lesser extent. Given this mutual attenuation, it appears that the associations observed at this locus represent one signal.

Despite the findings that the lead SNV (rs6453022; p.Pro284Gln) explains a substantial portion of the observed signal, the possibility of multiple contributing variants at this locus is supported by the fact that our previous MAGMA gene-based analyses performed by Ahmed *et al*. revealed that *ARHGEF28* gene-based analyses uncovered stronger associations than any individual variant associations (gene-based *P*=4.17x10^-11^ vs. *P*=1.97×10^-9^ for the most significant SNV/STR).^9^ Therefore, to further examine whether other variants within this locus may be classified as possibly causal, we performed joint fine-mapping of SNVs and STRs. These analyses revealed that all STRs and SNVs, including the two missense variants within this region, showed very low PIPs, with an STR (chr5:73778077:A_n_; *P*=4.54×10^-5^) receiving the highest PIP score (0.063) (**Table 1**). These findings support the lack of a single causal variant within this region. These results may also indicate shortcomings in fine-mapping tools for evaluating imputed STR associations.

Functional annotation of the lead STR variant (chr5:73778077:A_n_) using AlphaGenome revealed that this STR ranks in the top 1% of variants likely to impact gene expression and splice site usage in *ARHGEF28* in brain tissue (UBERON:0000955). The longer alternative alleles for this STR (17, 18, and 19 repeats) showed reduced *ARHGEF28* expression relative to the reference allele (16 repeats), with raw scores of -0.10, -0.17, and -0.80, respectively. In addition, this STR was predicted to influence splice site usage in the brain (raw score=0.77) (**Supplementary Table 3**). This STR is located within an Alu SINE element (AluSc; chr5:73777790-73778092). While this STR was not present in the brain TR-xQTL database, another GWAS-significant STR, chr5:73754888:ATTT_n_ (*P*=8.24×10^-8^), was strongly associated with DNA methylation at multiple CpG sites in the dorsolateral prefrontal cortex (DLPFC). These include cg23956565 (*P*=1.70×10^-16^), cg00119811 (*P*=6.79×10^-7^), cg27585641 (*P*=9.20×10^-6^) and cg21221899 (*P*=8.53×10^-5^), all located within or proximal to the *ARHGEF28* locus and overlap with multiple transcription factor binding sites (**Supplementary Table 4**). These mQTL associations suggest that the STR may modulate *ARHGEF28* gene regulation by altering methylation in brain tissues (**Supplementary Table 1**).

### 3.4. Investigation of haplotypes associated with metabolic hearing loss

Given that no single variant was identified as a clear driver of the metabolic ARHL signal on chromosome 5, it is possible that multiple variants within this locus act together to increase risk. To test this, we constructed a haplotype consisting of the two missense variants (rs6453022; p.Pro284Gln and rs7714670; p.Trp225Arg) and the STR with the highest PIP score (chr5:73778077:A_n_). Haplotype analyses revealed that only two STR-SNP combinations were significantly associated with metabolic ARHL. The haplotype carrying all three reference alleles [rs6453022 (C), rs7714670 (T), chr5:73778077 (A_16_)] was associated with protection against metabolic ARHL (*P*=3.3×10^-9^, standardized beta = -1.21). In contrast, the haplotype carrying alternative alleles at all three positions [rs6453022 (A), rs7714670 (A), chr5:73778077 (A_17_)] was associated with an increased risk of metabolic ARHL (*P*=3.07×10^-8^, standardized beta = 1.09). Other common haplotypes, including those containing only one missense variant, showed no significant association with metabolic ARHL (*P*>0.05). These results suggest that the haplotype containing both missense variants and the chr5:73778077:A_17_ STR variant is potentially responsible for the phenotypic variation observed for metabolic ARHL (**Table 2**).

**Table 2:** Analysis of common haplotypes within the *ARHGEF28* locus.

| CHR | Haploblock rs7714670_73778077:A<sub>n</sub>_rs6453022 | Haplotype frequency | Standardized Beta | *P* value |
| --- | --- | --- | --- | --- |
| 5 | T_A16_C | 0.46 | -1.21 | 3.30×10<sup>-9</sup> |
|  | **C_A17_A** | 0.45 | 1.09 | 3.07×10<sup>-8</sup> |
|  | T_A18_A | 0.05 | 0.48 | 0.29 |
|  | T_A19_C | 0.03 | -0.35 | 0.57 |

CHR: Chromosome. Alternative alleles are bolded

### 3.5. Investigation of variants nominally associated with metabolic and sensory hearing loss

Investigation of STRs nominally associated with metabolic hearing loss (*P*<1×10^-5^) uncovered 41 STRs in 9 independent loci. Three of these STRs exhibited stronger associations than SNVs within the same locus. First, an STR on chromosome 1 (chr1:90572148:GT_n_, *P*=1.96×10^-6^) mapped to *LINC02787* and is associated with multiple sQTL clusters for this gene (81746; *P*=2.91×10^-14^, 80954; *P*=2.13×10^-10^ and 78711; *P*=3.70×10^-7^), as well as being identified as an mQTL (cg09547692, *P*=1.39×10^-4^). Second, an STR on chromosome 2 (chr2:85268768:A_n_, *P*=4.09×10^-6^) mapped to *TCF7L1*, a transcription factor in the Wnt signaling pathway that is involved in cochlear development, particularly in hair cell differentiation and maturation.^44–47^ Third, an independent signal on chromosome 5 (chr5:158111773:A_n_, *P*=8.65×10^-7^), is located in a region lacking current gene annotation (**Figure 1, Supplementary Table 1**). While investigation of STRs that were nominally associated with estimates of the sensory component of ARHL identified 14 STRs, mapping to 6 independent loci (**Figure 1, Supplementary Table 2**), none of these STRs exhibited more significant associations than were observed for SNVs in the same loci.

### 3.6. Identification of STR expansion burden on sensory and metabolic hearing loss

Investigation of the cumulative burden of STR repeat expansions revealed that the presence of longer STR expansions was associated with increased risk of metabolic ARHL, with the effects of these associations increased for rare STRs. Individuals carrying STR variants that were ≥10 repeats longer than the reference and observed ≤ 5 times in the cohort showed a 223-fold higher effect (standardized beta = 0.67, FDR-adjusted *P*=0.022) compared to shorter and more frequent expansions (≥1 repeat longer than the reference, seen ≤100 times) (standardized beta = 0.003, FDR-adjusted *P*=0.019) (**Figure 2-A, Supplementary Table 5**). Strikingly, a similar pattern was observed for the sensory phenotype, but in the opposite direction, where long and rare expansions were significantly associated with a reduced risk of sensory ARHL (≥10 repeats longer than the reference, observed ≤5 times; standardized beta = −0.47, FDR-adjusted *P* = 2.34 × 10^-13^), compared to shorter and more frequent expansions (≥1 repeat longer than the reference, observed ≥100 times; standardized beta = −0.002, FDR-adjusted *P* = 2.2 × 10^-15^) (**Figure 2-B, Supplementary Table 5**). This highlights a clear distinction in the impact of STR length and frequency, with long and rare expansions having more pronounced effects on both phenotypes, although in opposite directions.

**Figure 2:**
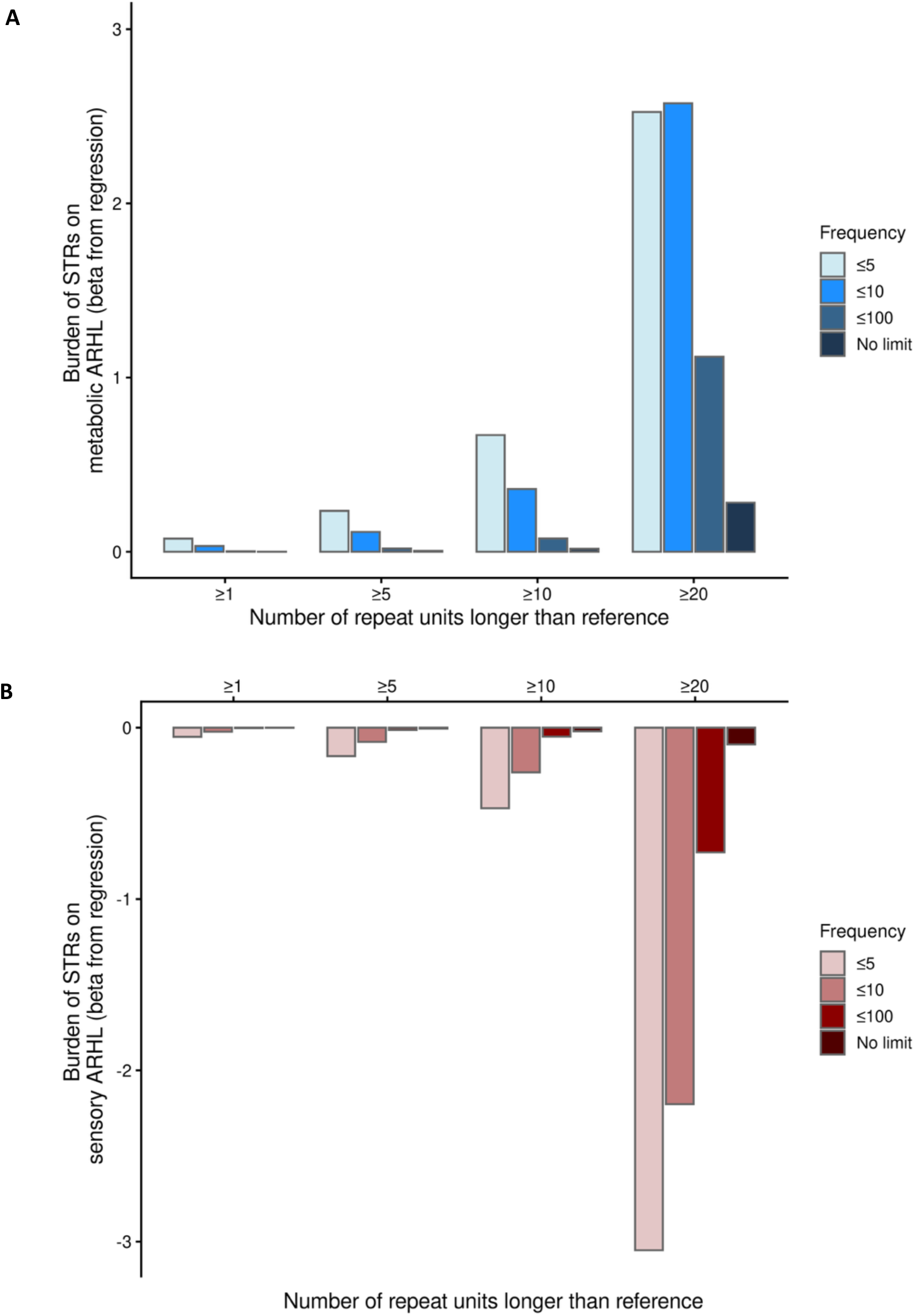
STR burden analysis showing beta coefficients from linear regression models testing the association between STR expansion length (at different allele frequency cutoffs) and **(A)** metabolic and **(B)** sensory hearing loss phenotypes of ARHL. STR: Short tandem repeat.

## 4. Discussion

In this first large-scale human genomics study investigating the role of STRs in ARHL, we demonstrate that STRs contribute to the genetic architecture of both metabolic and sensory ARHL. Beyond the variants identified through traditional SNV-based GWAS,^9^ STRs uncovered from the genome-wide scans performed in this study provide an additional layer of insight, representing an understudied yet important class of genetic variants. For the first time, we have shown that STRs may capture some of the missing heritability attributed to ARHL. Further, we have created a new resource of imputed STRs that can be included in future research using the CLSA dataset to perform similar studies for other human phenotypes.

Our heritability analysis revealed that STRs independently contribute to the total phenotypic variance observed for metabolic (6.16%) and sensory (3.75%) ARHL. These findings highlight the modest yet significant role of STRs in explaining the genetic variance of metabolic and sensory phenotypes of ARHL, with STRs playing a greater role in metabolic ARHL, a phenotype more substantially influenced by aging.^6^ These estimates are also comparable to STR heritability recently observed for other neurodegenerative disorders, including Parkinson’s disease (7% STR heritability) and Alzheimer’s disease (3% STR heritability).^48,30^ In line with the potentially more important role of STRs in metabolic ARHL, we observed that the lead variants in all nominally associated loci for sensory ARHL were SNVs. This contrasts with metabolic ARHL, where four independent signals were identified, with STRs as the lead signals.

STRs are known to influence the regulatory activity of genes involved in neuronal function through multiple mechanisms, including effects on splicing, gene expression, and DNA methylation of nearby loci.^48,49^ Such regulatory roles provide a biological rationale for their potential impact on sensory systems, including auditory pathways. In our STR-based GWAS, we identified a single locus reaching genome-wide significance. This locus mapped to *ARHGEF28*, a region which we have previously shown to be associated with metabolic ARHL through SNV-based GWAS.^9^ Conditional analyses, combined with fine-mapping and haplotype analyses, indicated that this signal may be driven by a haplotype, which contains both previously associated missense variants in *ARHGEF28*, along with an STR (chr5:73778077:A_17_) in an intron of this gene. This STR is located within an *Alu* unit, which is known to regulate gene expression post transcriptionally.^50^ Annotation with AlphaGenome predicted that this variant may result in decreased expression of *ARHGEF28* in diverse brain tissues, suggesting that this STR may have a functional role rather than only tagging the nearby missense SNVs. As both missense variants have previously been reported as benign (SIFT) and tolerated (PolyPhen) for all *ARHGEF28* transcripts,^9^ these findings suggest that the presence of both missense variants, along with the longer allele of the STR may act together to increase risk for metabolic ARHL. Further, results from the TR-xQTL atlas suggest that STRs in this region (chr5:73754888:ATTT_n_) significantly influence the methylation of the *ARHGEF28* gene, consistent with the well-established role of STRs affecting the methylation of over 100 genes.^17^ Notably, the associated STRs have A/T rich motifs, which are known to exhibit high rates of DNA polymerase slippage and increased susceptibility to somatic expansions.^51^ As metabolic ARHL is precipitated by aging, the effects of this STR may be further amplified by somatic expansions that occur during aging.

Beyond investigation of individual loci that are associated with metabolic and sensory ARHL, our burden analysis revealed that rare STR expansions of longer length are associated with increased risk of metabolic ARHL, consistent with observations in other age-related neurological disorders. This includes Alzheimer’s disease, where rare, expanded STR alleles contribute substantially to disease burden.^14^ Interestingly, the sensory phenotype of ARHL showed the opposite pattern, with rare expansions of longer length associated with reduced risk. Because longer STRs are more likely to expand during aging and metabolic ARHL is an age-related phenotype, the direction of effect we observe is biologically plausible. In contrast, sensory ARHL is less impacted by aging,^6^ suggesting that somatic expansions may play a smaller role in this phenotype. It is worth noting that the protective effect observed for the sensory phenotype may, in part, reflect a statistical artifact arising from the model used to estimate ARHL components.^6^ In individuals with substantial overall hearing loss, metabolic loss may obscure the sensory component, potentially biasing the estimated direction of effect. Instead, STRs contributing to sensory phenotype may act as xQTLs independent of age-related expansion. Together, these findings warrant further investigation to understand the molecular mechanisms underlying these associations.

Our study provides valuable insight into the role of STRs in the genetic architecture of ARHL subtypes; however, several limitations should be acknowledged. First, although the reference panel used for STR imputation includes individuals from diverse ancestries, the ARHL phenotype data analyzed in this study is predominantly composed of individuals of European ancestry. Second, the STR genotypes used in this study were imputed from SNVs in high LD. Therefore, STRs that are not in LD with nearby SNVs may have been missed. As a result, our analysis may not capture the full spectrum of STR variation. To overcome this limitation, future studies should incorporate STR genotypes obtained directly from PCR-free whole genome sequencing data rather than relying solely on imputation. Third, although our analysis focused on germline STR variation, future studies should also consider somatic repeat expansions, which are known to contribute to the pathogenesis of late-onset neurodegenerative disorders such as Huntington’s disease.^52^ Further, while the brain has been used as a proxy to examine gene expression patterns of relevance to the ear,^40^ there is currently no STR xQTL data available specifically for the inner ear, which limits our ability to directly assess the regulatory impact of STRs on gene expression in tissues from the peripheral auditory system. Finally, while this study lacks a formal replication cohort, we provide multiple layers of supporting evidence, including fine-mapping, functional annotation, and prior associations with ARHL phenotypes. Given the limited availability of STR data in independent cohorts, replication remains a challenge in the field, but our findings establish a foundation for future validation studies.

## 5. Conclusion

Our study highlights, for the first time, the non-redundant role of STRs in ARHL. By using imputed STR genotypes alongside accurately measured audiometric phenotypes of ARHL, we were able to estimate the independent contribution of STRs to the heritability of ARHL and uncover associations not captured by SNVs alone. Functional annotation revealed that STRs can influence both gene expression levels and alternative splicing of genes involved in ARHL. These findings not only reinforce the biological relevance of STRs in aging and hearing loss but also emphasize the need for future studies to integrate multiple classes of genetic variation, particularly for complex traits with incomplete SNV heritability. This can, in turn, advance our understanding of the genetic mechanisms underlying hearing loss and its shared biological pathways with ARHL and neurodegeneration. Finally, we created a new resource of imputed STRs for the CLSA dataset that is available for other researchers, enabling future studies to investigate the contribution of STRs to other complex phenotypes.

## Acknowledgements and funding

This research was made possible using the data/biospecimens collected by the Canadian Longitudinal Study on Aging (CLSA). Funding for the CLSA is provided by the Government of Canada through the Canadian Institutes of Health Research (CIHR) under grant reference: LSA 94473 and the Canada Foundation for Innovation, as well as the following provinces, Newfoundland, Nova Scotia, Quebec, Ontario, Manitoba, Alberta, and British Columbia. This research has been conducted using the CLSA Baseline Comprehensive Dataset Version 6.0 and Genome-wide Genetic Data Version 3.0 under Application Number 2104035. The CLSA is led by Drs. Parminder Raina, Christina Wolfson and Susan Kirkland. The opinions expressed in this manuscript are the author’s own and do not reflect the views of the Canadian Longitudinal Study on Aging. The authors gratefully acknowledge the time and commitment of the CLSA participants, without whom this research would not be possible.

This work was funded through a Natural Sciences and Engineering Research Council of Canada Discovery Grant (B.I.D.) (RGPIN-2022-04500), Canadian Institutes of Health Research Catalyst Grant (B.I.D/G.E.B.W.) (AFF-187272) and (in part) by the National Institutes of Health/National Institute on Deafness and Other Communication Disorders Clinical Research Center (P50 DC 000422) awarded to the Medical University of South Carolina and by the South Carolina Clinical and Translational Research (SCTR) Institute, with an academic home at the Medical University of South Carolina, NIH/NCATS Grant number UL1 TR001450 (J.R.D. and K.V.). Portions of this investigation were conducted in a facility constructed with support from Research Facilities Improvement Program Grant Number C06 RR14516 from the NIH/NCRR (J.R.D. and K.V.). B.I.D. is supported by a CIHR Tier 2 Canada Research Chair in *Pharmacogenomics and Precision Medicine* (CRC-2019-00040/CRC-2023-00351). G.E.B.W. is supported by an NSERC Tier 2 Canada Research Chair in *Neurogenomics* (CRC-2019-145/CRC-2025-00075) and an NSERC Discovery Grant Program (RGPIN-2022-04509). S.A. was supported through a Research Manitoba Studentship and the NSERC-CREATE Visual and Automated Disease Analytics Graduate Training Program at the University of Manitoba.

## 6. Author contributions

S.A. performed the discovery analyses and wrote the manuscript. K.I.V. and J.R.D. developed the phenotyping approach and provided expertise on ARHL in humans. G.E.B provided expertise relating to STRs. B.I.D. and G.E.B conceived of the project. B.I.D supervised the project. All authors contributed to editing the manuscript.

## 8. Data availability

Data, including imputed STR genotypes, are available from the Canadian Longitudinal Study on Aging (www.clsa-elcv.ca) for researchers who meet the criteria for access to de-identified CLSA data in addition to imputed STRs. Scripts that were used in this study are available on https://github.com/Drogemoller-Lab/STR

---

**Supplementary Figure 1:**
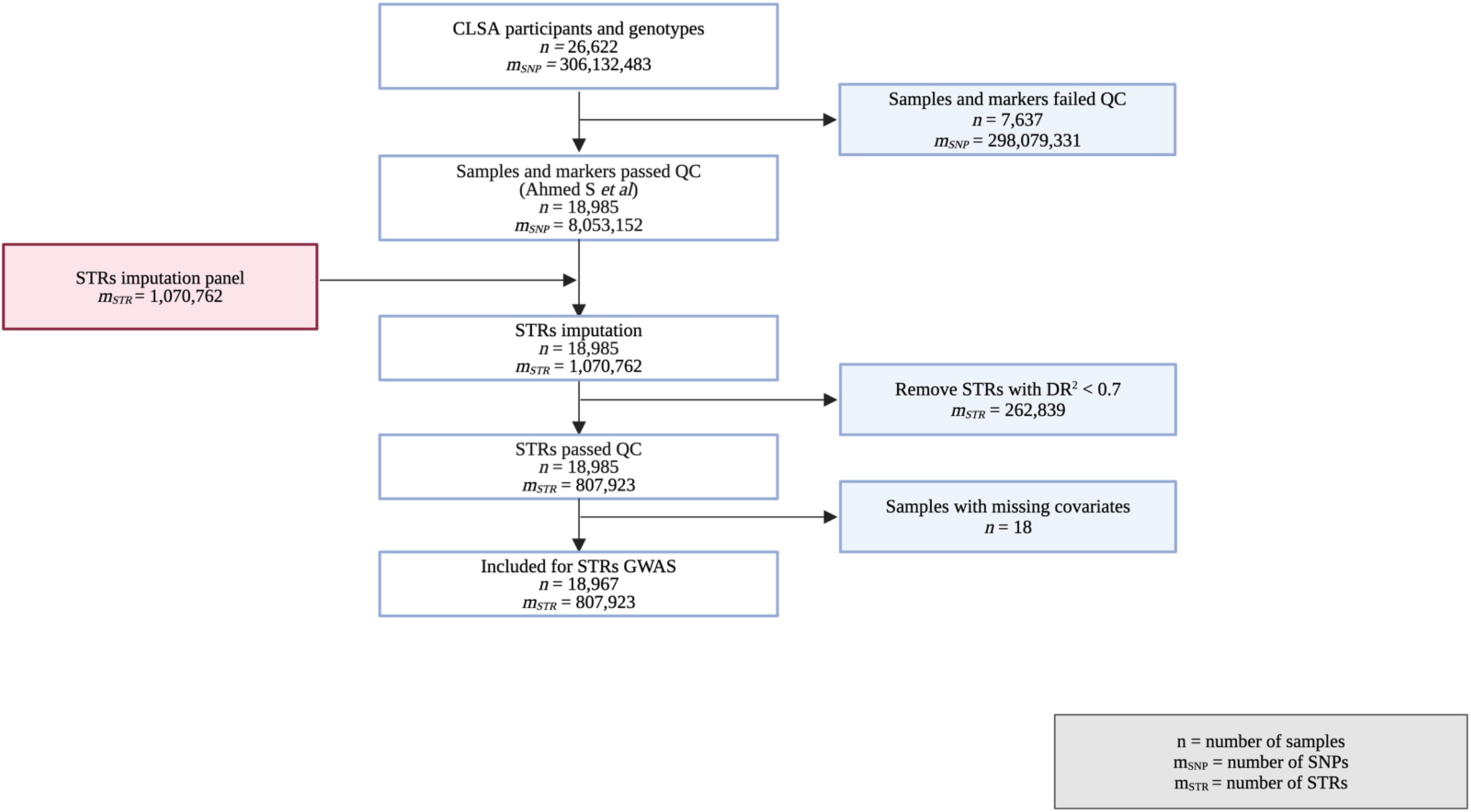
Flow diagram illustrating the QC steps for both samples and markers in the study. CLSA: Canadian Longitudinal Study on Aging, GWAS: Genome-wide association study, STR: Short Tandem Repeats, QC: Quality Control, DR^2^: Dosage R^2^.

**Supplementary Figure 2:**
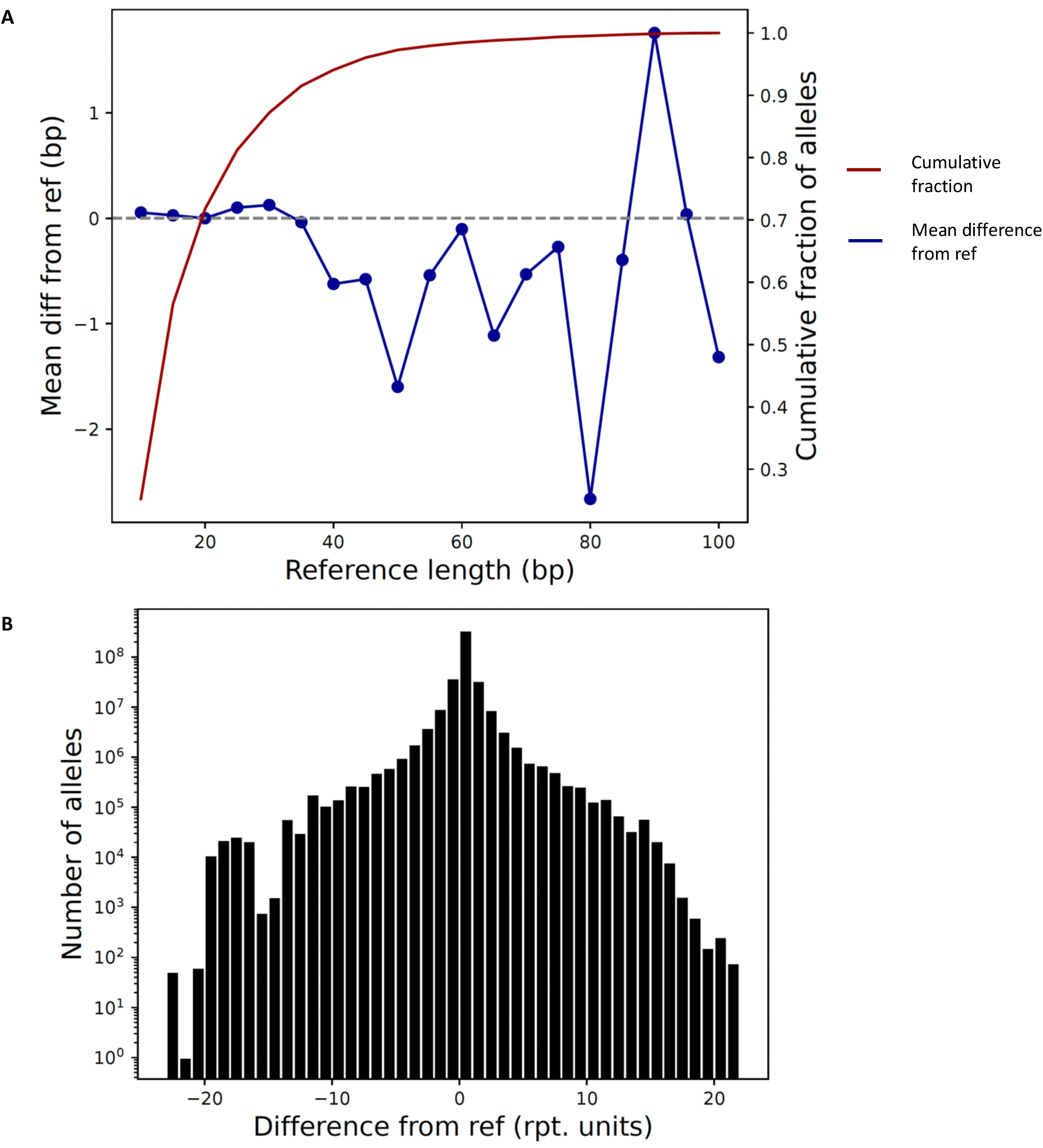
Imputation quality of the CLSA cohort using the STR reference panel. (data shown from chromosome 22). **(A)** Plot of STR reference length versus the mean difference in length from the reference shows slightly reduced imputation quality with increasing repeat length. The red line indicates the cumulative fraction of STRs below each reference length. **(B)** Histogram of the difference in length between imputed alleles and the reference, showing minimal deviation and suggesting high overall imputation accuracy. Ref: Reference, Rpt: Repeats, CLSA: Canadian Longitudinal Study on Aging, STR: Short Tandem Repeats.

